# Drug-Coated Devices, Wound Healing, and Mortality After Endovascular Therapy for Chronic Limb-Threatening Ischemia

**DOI:** 10.64898/2026.07.14.26358110

**Authors:** Yonggu Lee, Alexander D Rodway, Gary D Maytham, Nikolaos Ntagiantas, Ivan Walton, Felipe Pazos-Casal, Charlotte Allan, Marianne Brodmann, Oliver Schlager, Jenny Harris, Christian Heiss

**Author notes:** **Corresponding author** Professor Christian Heiss Dr. med, FRCP, FESC, FRSM School of Medicine, Faculty of Health and Medical Sciences, University of Surrey 30 Priestley Road, Guildford GU2 7YH, UK.

## Abstract

**Background:** The clinical benefit and safety of drug-coated devices in chronic limb-threatening ischemia remain debated, particularly after recent randomized evidence questioning paclitaxel-coated technologies. We evaluated wound healing, limb outcomes, and mortality after infrainguinal endovascular therapy with uncoated, paclitaxel-coated, and sirolimus-coated devices.

**Methods:** Consecutive patients with chronic limb-threatening ischemia undergoing successful infrainguinal endovascular therapy in a prospective single-center service evaluation were analyzed. The primary exposure was use of any drug-coated device during the index procedure. Inverse probability of treatment weighting and multivariable Cox models were used to adjust for baseline differences. Exploratory analyses compared paclitaxel-coated, sirolimus-coated, and uncoated devices.

**Results:** Among 341 patients, 244 (71.6%) received at least one drug-coated device. After weighting, drug-coated device use was associated with more frequent wound healing, whereas major amputation, clinically driven target lesion revascularization, major adverse limb events, and death did not differ significantly between groups. In weighted multivariable models, drug-coated device use remained associated with wound healing (HR, 1.86; 95% CI, 1.14–3.02), but not with mortality or major limb events. Exploratory drug-specific analyses suggested the highest wound-healing rates among patients treated with sirolimus-coated devices, while mortality was comparable between paclitaxel-coated and uncoated devices.

**Conclusion:** In this real-world cohort of patients with chronic limb-threatening ischemia undergoing infrainguinal endovascular therapy, drug-coated device use was not associated with increased adjusted 1-year mortality and was associated with improved wound healing. Exploratory analyses suggested favourable wound-healing outcomes with sirolimus-coated balloons, with a lower observed mortality signal that warrants confirmation in larger comparative studies.

**What is Known:**

- Drug-coated devices are widely used in endovascular therapy to prevent restenosis, but their clinical benefit in patients with chronic limb-threatening ischemia remains uncertain.
- Recent randomized evidence has questioned the effectiveness of paclitaxel-coated devices in chronic limb-threatening ischemia and has raised ongoing concerns regarding long-term mortality.

**What the Study Adds:**

- In a real-world cohort of patients with chronic limb-threatening ischemia undergoing infrainguinal endovascular therapy, drug-coated device use was not associated with increased adjusted 1-year mortality and was associated with improved wound healing.
- Wound healing may be a clinically sensitive, patient-relevant endpoint for evaluating endovascular strategies in CLTI.
- Exploratory drug-specific analyses suggested favourable outcomes with sirolimus-coated devices, supporting dedicated comparative studies of limus-based technologies in CLTI.

## Introduction

Chronic limb-threatening ischemia (CLTI) is the most severe form of peripheral arterial disease (PAD) and carries substantial risks of major amputation and mortality. Antiproliferative drug-coated devices are widely used to mitigate restenosis and improve patency following endovascular therapy (EVT).^1,2^ However, recent large-scale randomized evidence has challenged this practice. In SWEDEPAD, paclitaxel-coated devices did not reduce major amputation or repeat revascularization in patients with CLTI at 5 years.^3^ Among claudicants, the same trial showed no improvement in quality of life and an increased risk of 5-year (but not 7-year) mortality, raising concerns regarding the long-term safety of paclitaxel-coated devices.^4^

Despite these findings and due to solid RCT evidence on their positive effects on patency, drug-coated devices remain the mainstay of EVT in patients with CLTI, while debate on their efficacy and safety continues. Several limitations of SWEDEPAD-including its pragmatic design, device heterogeneity, and lack of systematic control over drug dose and delivery-have been highlighted.^3^ Moreover, the one-year reduction in repeat revascularization observed with the paclitaxel-coated devices remains clinically relevant because wound healing typically occurs within 2-4 months,^5^ and even short-term patency gains may translate into meaningful limb outcomes. The absence of a consistent mortality signal across follow-up has further contributed to uncertainty.^4^ In addition, sirolimus-coated devices have recently been introduced into EVT, with promising early results that may influence contemporary practice.^6–8^ However, major amputation and repeat revascularization may incompletely capture clinically meaningful early benefit in CLTI, where sustained perfusion during the first months after revascularization may be critical for tissue repair. Wound healing is therefore a patient-relevant endpoint that may be particularly sensitive to early differences in vessel patency and recoil.

Therefore, this study aimed, in real-world patients with CLTI; (a) to determine whether drug-coated device use is associated with adverse clinical outcomes, (b) to evaluate whether outcomes differ according to antiproliferative drug type; and (c) to assess whether any observed differences between drug-coated and uncoated devices are driven by specific device platforms, including drug-coated balloons (DCB) and drug-eluting stents (DES).

## Methods

### Patients and data

From August 2018 to August 2025, consecutive patients referred to the vascular unit of the East Surrey Hospital with suspected PAD who subsequently underwent EVT were prospectively included in a rolling service evaluation. East Surrey Hospital is a secondary-care general hospital within Surrey and Sussex NHS Trust, located in Redhill, Surrey, UK, serving as the principal acute care provider for East Surrey and north-east West Sussex. The hospital operates a regional vascular unit supported by the vascular hub at St George’s University Hospitals NHS Foundation Trust, London, and by the Royal Sussex County Hospital, Brighton, the arterial center for the Sussex Vascular Network.

Patients requiring urgent EVT, those with body mass index ≥35 kg/m^2^, and those with advanced kidney disease defined as an estimated glomerular filtration rate (eGFR) <30 mL/min/1.72m^2^, were transferred to the vascular hubs and not included in the service evaluation. Asymptomatic patients (Fontaine I), those referred for claudication alone (Fontaine II), those who did not undergo EVT, and those with unsuccessful EVT, isolated iliac artery interventions, covered stent implantation, or missing clinical outcomes were excluded from the analyses. The study adhered to the Declaration of Helsinki; written informed consent was waived because data were collected as part of an NHS service evaluation (Audit No. 3352).

Demographic characteristics, comorbidities, medications, and laboratory results were extracted from electronic hospital records. Missing or unclear information was verified through face-to-face interviews in the day-care unit prior to EVT.

#### Diagnosis, procedure and outcome definitions

PAD was diagnosed as an estimated ankle-brachial pressure index (eABPI) ≤0.9. The eABPI was estimated using the Doppler-waveform-based acceleration index, as previously described.^9^ Clinical staging followed the Fontaine Classification. CLTI was defined as rest pain (Fontaine III) or ulcer/gangrene on the foot ipsilateral to the arterial lesions (Fontaine IV). Lower extremity ultrasonography with Doppler measurements were obtained in all patients before EVT, while lower extremity computerized tomography was selectively performed at the consultant’s discretion.

All EVTs were performed in the day-care unit by three senior interventionalists, aiming to establish inline flow to the wound site through at least one artery. Patients received dual antiplatelet therapy with aspirin and clopidogrel for 1-3 months post-procedure. Antegrade ipsilateral common femoral artery access under ultrasound guidance using a 5–7F sheath was the default approach; when unsuitable, a cross-over approach was used. A 5,000 IU of heparin was administered during EVT. Target lesions were predilated with plain balloons (PBs) and treated with paclitaxel-coated balloons (PCB; Passeo, Biotronik), sirolimus-coated balloon (SCB; Selution, Cordis), paclitaxel-eluting stents (PES; Zilver PTX, Cook; Eluvia, Boston Scientific), or bare nitinol stents (BNS; Supera, Abbott; E-Luminexx, Bard). Rotational atherectomy (Jetstream, Boston Scientific) was used for heavily calcified lesions. Haemostasis was achieved using a closure device (Angioseal, St. Jude Medical).

EVT success was defined as residual stenosis <30% in the target lesion. Clinical events were assessed through either telephone interviews or in-person vascular clinic visits. Wound healing was defined as complete epithelialization of the index wound without drainage, infection, or the need for further wound care and was documented by the respective wound care specialists. Major amputation was defined as amputation at or above the ankle. Clinically driven target lesion revascularization (cdTLR) was defined as repeat revascularization performed within the index lesion because of recurrent or worsening symptoms. Major adverse limb events (MALE) were defined as a composite of major amputation and cdTLR.^10,11^

#### Statistical analysis

Patients were categorized into two groups: the drug-coated device group included infrainguinal EVTs with PCB, SCB, or PES, and the uncoated device group included EVT with BNS or PB angioplasty only. The primary exposure represented use of any drug-coated technology as part of the index EVT strategy, rather than assignment to a single device platform. Baseline characteristics were compared using Student’s t-tests for normally distributed continuous variables, Mann–Whitney U-tests for non-normally distributed continuous variables, χ² tests for categorical variables, and Fisher’s exact tests when any cell count was ≤5.

To balance baseline difference, inverse probability of treatment weighting (IPTW) was applied. The IPTW model included age, sex, current smoking, WIfI stage components, heel ulcer, eABPI, rest pain, Fontaine classification, diabetes, hypertension, coronary artery disease, heart failure, atrial fibrillation, stroke, antiplatelet use, eGFR, haemoglobin A1c, lipid profiles, and lesion characteristics, including numbers of lesions, total lesion length and presence of chronic total occlusion (CTO). Balance after IPTW was assessed using standardized mean differences (SMDs); SMD <0.1 was considered full balance and SMD >0.2 indicated substantial imbalance between groups.

IPTW-applied Kaplan-Meier curve analyses with log-rank tests were used to compare clinical outcomes. IPTW-applied multivariable Cox proportional hazards (CPH) models were then used to identify the association between drug-coated device use and clinical outcomes. Variables with SMD >0.1 after IPTW were included as covariates in the multivariable models to further adjust residual confounding, unless collinearity with related variables precluded inclusion.

For outcomes associated with drug-coated device use, stratified analyses were performed according to rest pain, wound, ischemia, infection, WIfI stage, diabetes, and lesion location to assess potential treatment interactions. A subgroup analysis was performed among patients treated exclusively with single type of drug-coated or uncoated devices to compare clinical outcomes according to drug type. Multinomial IPTW was applied to balance baseline differences across sirolimus-coated, paclitaxel-coated, and uncoated devices, followed by multivariable CPH models to adjust for residual differences. Additional subgroup analyses were performed in patients treated without stent implantation and in those treated without DCB application to identify whether specific device platforms drove differences in clinical outcomes. In both subgroups, IPTW and multivariable CPH models were applied to balance baseline characteristics and evaluate outcome associations.

All statistical analyses were performed using R-4.2.2 (R Core Team, R Foundation for Statistical Computing, Vienna, Austria) in RStudio-2023.12.1 (Build 402; RStudio Team, BPC, Boston, MA, USA). A P value <0.05 was considered significant.

## Results

A total of 547 consecutive patients were referred for EVT. Among 455 consecutive patients with CLTI who underwent EVT, 341 patients with available clinical outcomes after infrainguinal EVT were analysed (Figure 1). The uncoated device group comprised 97 patients (28.4%), of whom 80 (82.4%) underwent PB angioplasty only. The drug-coated device group comprised 244 patients; DCB was the most frequently used device (n=124; 36.7% of all patients and 51.2% of the drug-coated device group).

**Figure 1.**
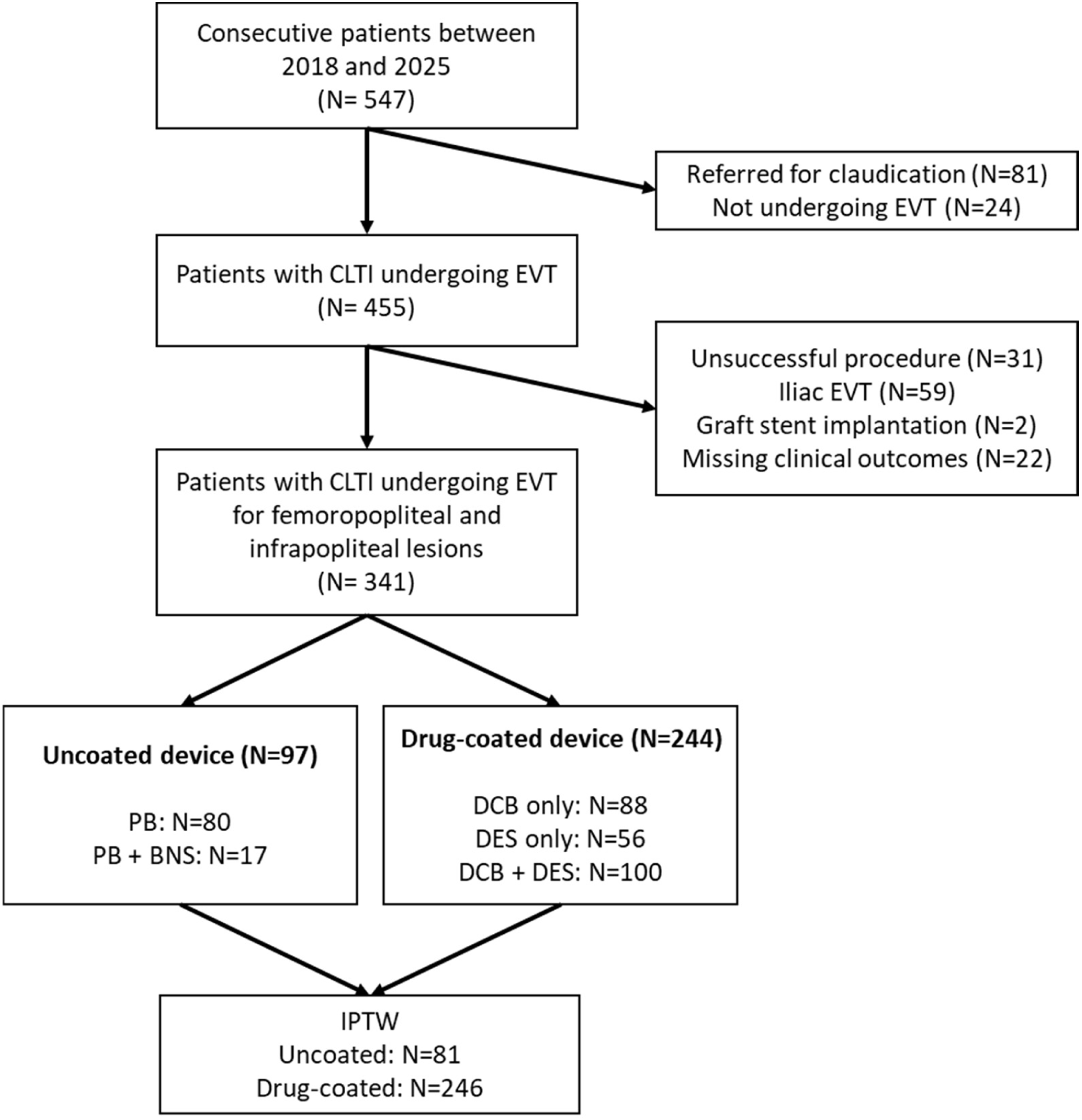
Patient selection process. Among 547 consecutive patients referred for EVT between 2018 and 2025, 341 patients with CLTI who underwent successful EVT for infrainguinal lesions were included in the analysis. IPTW was applied to balance baseline characteristics between the uncoated and drug-coated device groups. *All DESs were paclitaxel-eluting stents. CLTI, chronic limb-threatening ischemia; EVT, endovascular therapy; IPTW, inverse probability of treatment weighting; PB, plain balloon; BNS, bare-nitinol stent; DCB, drug-coated balloon; DES, drug-eluting stent.

Baseline characteristics are summarized in Table 1. After IPTW, the sum of weights corresponded to 81 and 246 patients in the uncoated and drug-coated groups, and most variables were well balanced (Figure S1). However, the wound component of the WIfI stage remained lower, whereas non-high-density lipoprotein cholesterol (non-HDL-C) levels and lesion complexity including total lesion length and the number of lesion levels involved, remained higher in the drug-coated device group.

**Table 1.** Baseline characteristics of patients in both unweighted cohort and IPTW-applied cohort.

| n | Unweighted cohort |  | p | IPTW-applied cohort |  | p | SMD |
| --- | --- | --- | --- | --- | --- | --- | --- |
|  | Uncoated | Drug-coated |  | Uncoated | Drug-coated |  |  |
|  | 97 | 244 |  | 81 | 246 |  |  |
| Age (years) | 75.1 ± 11.7 | 74.3 ± 10.9 | 0.598 | 75.7 ± 11.1 | 74.7 ± 10.9 | 0.501 | 0.093 |
| Male sex | 67 (69.1) | 147 (60.2) | 0.130 | 55.6 (69.1) | 154.7 (62.9) | 0.372 | 0.131 |
| Current smoker | 10 (10.3) | 57 (23.4) | 0.007 | 14.6 (18.1) | 47.8 (19.4) | 0.849 | 0.034 |
| IMD (decile) | 7.1 ± 2.2 | 7.1 ± 2.2 | 0.907 | 7.1 ± 2.2 | 7.2 ± 2.2 | 0.892 | 0.021 |
| Disease severity |  |  |  |  |  |  |  |
| Pre-EVT ABPI | 0.51 ± 0.28 | 0.49 ± 0.21 | 0.582 | 0.50 ± 0.24 | 0.51 ± 0.21 | 0.923 | 0.013 |
| Rest pain | 73 (75.3) | 182 (74.6) | 0.898 | 62.6 (77.7) | 183.5 (74.6) | 0.609 | 0.073 |
| Fontaine class III | 5 (5.2) | 41 (17.2) | 0.004 | 10 (11.8) | 33 (13.5) | 0.795 | 0.050 |
| IV | 92 (94.8) | 202 (82.8) |  | 71 (88.2) | 213 (86.5) |  |  |
| Heel ulcer | 7 (7.2) | 24 (9.8) | 0.522 | 5 (6.1) | 21 (8.4) | 0.522 | 0.086 |
| WIFI score |  |  | 0.027 |  |  | 0.501 | 0.150 |
| 1 | 1 (1.0) | 12 (4.9) |  | 1 (0.8) | 12 (4.9) |  |  |
| 2 | 10 (10.3) | 50 (20.5) |  | 14 (16.8) | 44 (17.8) |  |  |
| 3 | 34 (35.1) | 81 (33.2) |  | 26 (32.5) | 76 (30.9) |  |  |
| 4 | 52 (53.6) | 101 (41.4) |  | 40 (49.9) | 114 (46.5) |  |  |
| Wound component |  |  | <0.001 |  |  | 0.089 | 0.155 |
| 0 | 5 (5.2) | 36 (14.8) |  | 10 (11.8) | 28 (11.4) |  |  |
| 1 | 17 (17.5) | 87 (35.7) |  | 13 (16.3) | 79 (32.2) |  |  |
| 2 | 54 (55.7) | 95 (38.9) |  | 48 (59.0) | 103 (41.8) |  |  |
| 3 | 21 (21.6) | 26 (10.7) |  | 10 (12.9) | 36 (14.6) |  |  |
| Ischemia component |  |  | 0.032 |  |  | 0.803 | 0.080 |
| 0 | 6 (6.2) | 8 (3.3) |  | 3 (4.0) | 14 (5.5) |  |  |
| 1 | 38 (39.2) | 62 (25.4) |  | 26 (32.7) | 66 (26.9) |  |  |
| 2 | 31 (32.0) | 104 (42.6) |  | 33 (40.8) | 107 (43.6) |  |  |
| 3 | 22 (22.7) | 70 (28.7) |  | 18 (22.5) | 59 (24.0) |  |  |
| Infection component |  |  | 0.022 |  |  | 0.382 | 0.077 |
| 0 | 21 (21.6) | 95 (38.9) |  | 21 (26.6) | 84 (34.2) |  |  |
| 1 | 44 (45.4) | 91 (37.3) |  | 37 (45.7) | 96 (39.1) |  |  |
| 2 | 31 (32.0) | 55 (22.5) |  | 22 (27.2) | 59 (24.0) |  |  |
| 3 | 1 (1.0) | 3 (1.2) |  | 0 (0.0) | 7 (2.7) |  |  |
| Comorbidity |  |  |  |  |  |  |  |
| Diabetes | 69 (71.1) | 155 (63.5) | 0.183 | 57 (70.6) | 165 (67.0) | 0.616 | 0.077 |
| Diabetes on insulin | 36 (37.1) | 82 (33.6) | 0.540 | 27 (34.0) | 87 (35.5) | 0.834 | 0.031 |
| Hypertension | 79 (81.4) | 201 (82.4) | 0.840 | 69 (85.4) | 206 (83.6) | 0.720 | 0.050 |
| Coronary artery disease | 46 (47.4) | 95 (38.9) | 0.153 | 36 (44.3) | 107 (43.3) | 0.896 | 0.020 |
| Heart failure | 23 (23.7) | 50 (20.5) | 0.514 | 20 (25.3) | 58 (23.7) | 0.808 | 0.037 |
| Atrial fibrillation | 31 (32.0) | 56 (23.0) | 0.087 | 20 (24.9) | 60 (24.4) | 0.940 | 0.011 |
| Deep vein thrombosis | 4 (4.1) | 8 (3.3) | 0.703 | 3 (3.6) | 6 (2.5) | 0.582 | 0.063 |
| Stroke | 15 (15.5) | 40 (16.4) | 0.834 | 11 (14.2) | 40 (16.1) | 0.731 | 0.054 |
| Medications |  |  |  |  |  |  |  |
| Aspirin | 45 (46.4) | 97 (39.8) | 0.263 | 37 (46.4) | 105 (42.7) | 0.616 | 0.076 |
| Clopidogrel | 37 (38.1) | 94 (38.5) | 0.948 | 35 (42.9) | 94 (38.3) | 0.545 | 0.094 |
| Oral anticoagulation | 33 (34.0) | 65 (26.6) | 0.176 | 22 (27.6) | 69 (28.2) | 0.928 | 0.013 |
| Statin | 66 (68.0) | 176 (72.1) | 0.454 | 57 (70.1) | 180 (73.2) | 0.644 | 0.068 |
| RAS blocker | 57 (58.8) | 138 (56.6) | 0.711 | 45 (55.9) | 144 (58.6) | 0.730 | 0.053 |
| Beta blocker | 40 (41.2) | 104 (42.6) | 0.816 | 30 (37.5) | 110 (44.8) | 0.321 | 0.148 |
| Calcium channel blocker | 23 (23.7) | 55 (22.5) | 0.817 | 19 (23.9) | 54 (21.9) | 0.743 | 0.048 |
| Laboratory data |  |  |  |  |  |  |  |
| eGFR (mL/min/1.72 m<sup>2</sup>) | 62.3 ± 21.4 | 66.6 ± 23.4 | 0.102 | 61.8 ± 20.5 | 64.2 ± 23.2 | 0.422 | 0.110 |
| HbA1c (mmol/mol) | 58.4 ± 19.1 | 56.8 ± 20.5 | 0.480 | 56.7 ± 19.8 | 57.3 ± 20.3 | 0.858 | 0.026 |
| CRP (g/L) | 28 (13, 49) | 30 (14, 51) | 0.631 | 27 (12, 46) | 32 (14, 55) | 0.420 | 0.078 |
| Total cholesterol (mmol/L) | 4.2 ± 1.1 | 4.6 ± 1.3 | 0.022 | 4.2 ± 1.1 | 4.4 ± 1.2 | 0.182 | 0.183 |
| Non-HDL cholesterol (mmol/L) | 2.9 ± 1.1 | 3.3 ± 1.3 | 0.009 | 2.9 ± 1.0 | 3.2 ± 1.2 | 0.103 | 0.217 |
| Triglyceride (mmol/L) | 1.8 (1.1, 2.5) | 2.1 (1.3, 3.0) | 0.002 | 1.8 (1.1, 2.5) | 1.9 (1.2, 2.8) | 0.173 | 0.251 |
| LDL cholesterol (mmol/L) | 2.1 ± 0.9 | 2.2 ± 0.9 | 0.264 | 2.1 ± 0.8 | 2.2 ± 0.9 | 0.405 | 0.111 |
| HDL cholesterol (mmol/L) | 1.3 ± 0.3 | 1.3 ± 0.4 | 0.445 | 1.3 ± 0.3 | 1.3 ± 0.4 | 0.560 | 0.086 |
| Lesion characteristics |  |  |  |  |  |  |  |
| Levels of lesions |  |  | <0.001 |  |  | 0.156 | 0.196 |
| 1 | 66 (68.0) | 87 (35.7) |  | 47 (58.4) | 110 (44.5) |  |  |
| 2 | 22 (22.7) | 110 (45.1) |  | 21 (25.6) | 95 (38.8) |  |  |
| 3 | 9 (9.3) | 47 (19.3) |  | 13 (16.0) | 41 (16.7) |  |  |
| Femoropopliteal lesions | 43 (44.3) | 211 (86.5) | <0.001 | 55 (68.5) | 183 (74.2) | 0.364 | 0.127 |
| Infrapopliteal lesions | 76 (78.4) | 130 (53.3) | <0.001 | 50 (61.9) | 148 (59.9) | 0.799 | 0.041 |
| Lesion length (mm) | 165 (100, 300) | 240 (120, 350) | 0.012 | 165 (80, 304) | 220 (118, 350) | 0.186 | 0.179 |
| CTO | 85 (87.6) | 216 (88.5) | 0.817 | 73 (90.6) | 215 (87.5) | 0.453 | 0.098 |
| Femoropopliteal CTO | 33 (34.0) | 180 (73.8) | <0.001 | 47 (57.9) | 151 (61.5) | 0.610 | 0.075 |
| Infrapopliteal CTO | 68 (70.1) | 147 (60.2) | 0.090 | 49 (59.3) | 156 (63.4) | 0.590 | 0.084 |

SMD; Standardized mean difference; IMD, Index of multiple deprivation; EVT, endovascular; ABPI, Ankle-brachial pressure index; RAS, Renin-Angiotensin system; eGFR, estimated glomerular filtration rate; HbA1c, Hemoglobin A1c; CRP, C-reactive protein; HDL, High-density lipoprotein; LDL, Low-density lipoprotein; CTO, chronic total occlusion
The data were shown in the mean ± SD or N (%).
The median (Interquartile range) was presented when data were skewed

Wound healing was achieved in 182 (53.4%) patients, while major amputation, cdTLR, MALE and death occurred in 23 (6.7%), 30 (8.8%), 47 (13.8%) and 54 (15.8%) patients, respectively. The numbers of clinical events are presented in Table 2. Unweighted Kaplan–Meier analyses showed that wound healing was higher and mortality was lower in the drug-coated device group than in the uncoated device group (Figure S2). IPTW-applied Kaplan–Meier analyses showed a higher wound healing rate in the drug-coated device group, whereas the incidences of major amputation, cdTLR, MALE, and death did not differ between groups (Figure 2). In IPTW-applied multivariable CPH models, drug-coated device use was associated with a higher probability of wound healing, but not with higher risks of other clinical outcomes (Table 2). In stratified analyses, association between drug-coated devices and better wound healing was consistent across strata, although the association appeared slightly stronger in patients with rest pain, wound scores ≥2 or WIfI stage 4 (Figure S3).

**Figure 2.**
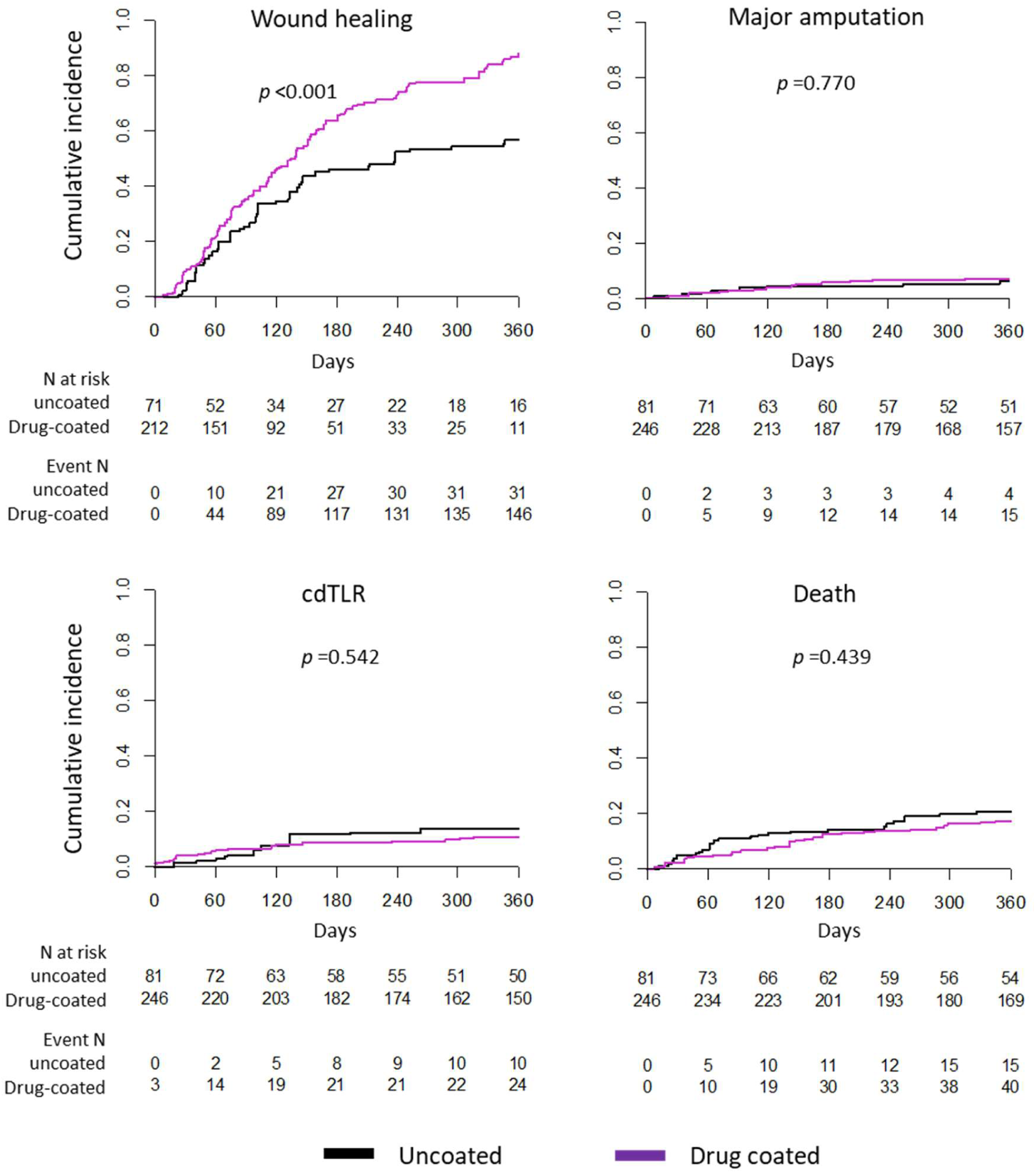
IPTW-applied cumulative incidences of clinical events according to the use of drug-coated devices. In the IPTW-applied cohort, the cumulative incidence of wound healing was significantly higher in the drug-coated device group than in the uncoated device group, whereas the incidences of major amputation, cdTLR, and death did not differ significantly between the groups. IPTW, inverse probability of treatment weighting; cdTLR, clinically-driven target lesion revascularization.

**Table 2.** Association of the use of drug-coated devices with clinical outcomes in the IPTW-applied cohort.

|  | Unweighted N |  | Unweighted event N |  | p-value* | Weighted univariable models |  |  | Weighted multivariable models |  |  |
| --- | --- | --- | --- | --- | --- | --- | --- | --- | --- | --- | --- |
|  | Uncoated | Drug-coated | Uncoated | Drug-coated |  | HR | 95% CI | p-value | HR | 95% CI | p-value |
| Wound healing | 92 | 201 | 42 (45.7%) | 140 (69.7%) | <0.001 | 1.99 | 1.20 - 3.32 | 0.008 | 1.86 | 1.14 - 3.02 | 0.012 |
| Major Amputation | 97 | 244 | 8 (8.2%) | 15 (6.1%) | 0.485 | 1.18 | 0.48 - 3.04 | 0.733 | 1.26 | 0.48 - 3.26 | 0.641 |
| cdTLR | 97 | 244 | 11 (11.3%) | 19 (7.8%) | 0.296 | 0.79 | 0.34 - 1.87 | 0.595 | 0.87 | 0.36 - 2.10 | 0.750 |
| MALE | 97 | 244 | 18 (18.6%) | 29 (11.9%) | 0.107 | 0.84 | 0.42 - 1.69 | 0.622 | 0.95 | 0.46 - 2.00 | 0.902 |
| Death | 97 | 244 | 23 (23.7%) | 31 (12.7%) | 0.012 | 0.79 | 0.44 - 1.43 | 0.440 | 0.90 | 0.47 - 1.71 | 0.744 |

All univariate and multivariate models were constructed in the balanced cohort using IPTW.
The weighted multivariable models additionally adjusted the variables with SMD >0.1 (sex, Wifl wound grade, non-HDL cholesterol, total lesion length, number of lesion levels, presence of femoropopliteal lesions).
\* $\chi^2$ test

Among patients treated with a single device type, 150 received paclitaxel-coated devices, 41 received sirolimus-coated devices, and 95 received uncoated devices. After multinomial IPTW, most baseline variables were balanced among the three groups, although WIfI wound scores remained lower and non-HDL-C levels and lesion complexity remained higher in the sirolimus- and paclitaxel-coated device groups than in the uncoated device group (Table S1, Figure S4). IPTW-applied Kaplan-Meier analyses showed that wound healing occurred most frequently in the sirolimus-coated device group (83.3%) and least frequently in the uncoated device group (42.2%; Figure 3). Mortality did not significantly differ among the three groups, although it was numerically the lowest in the sirolimus-coated device group, compared with the paclitaxel-coated and uncoated device group (9.7% vs. 18.1% vs. 21.9%). Other clinical outcomes were similar across groups.

**Figure 3.**
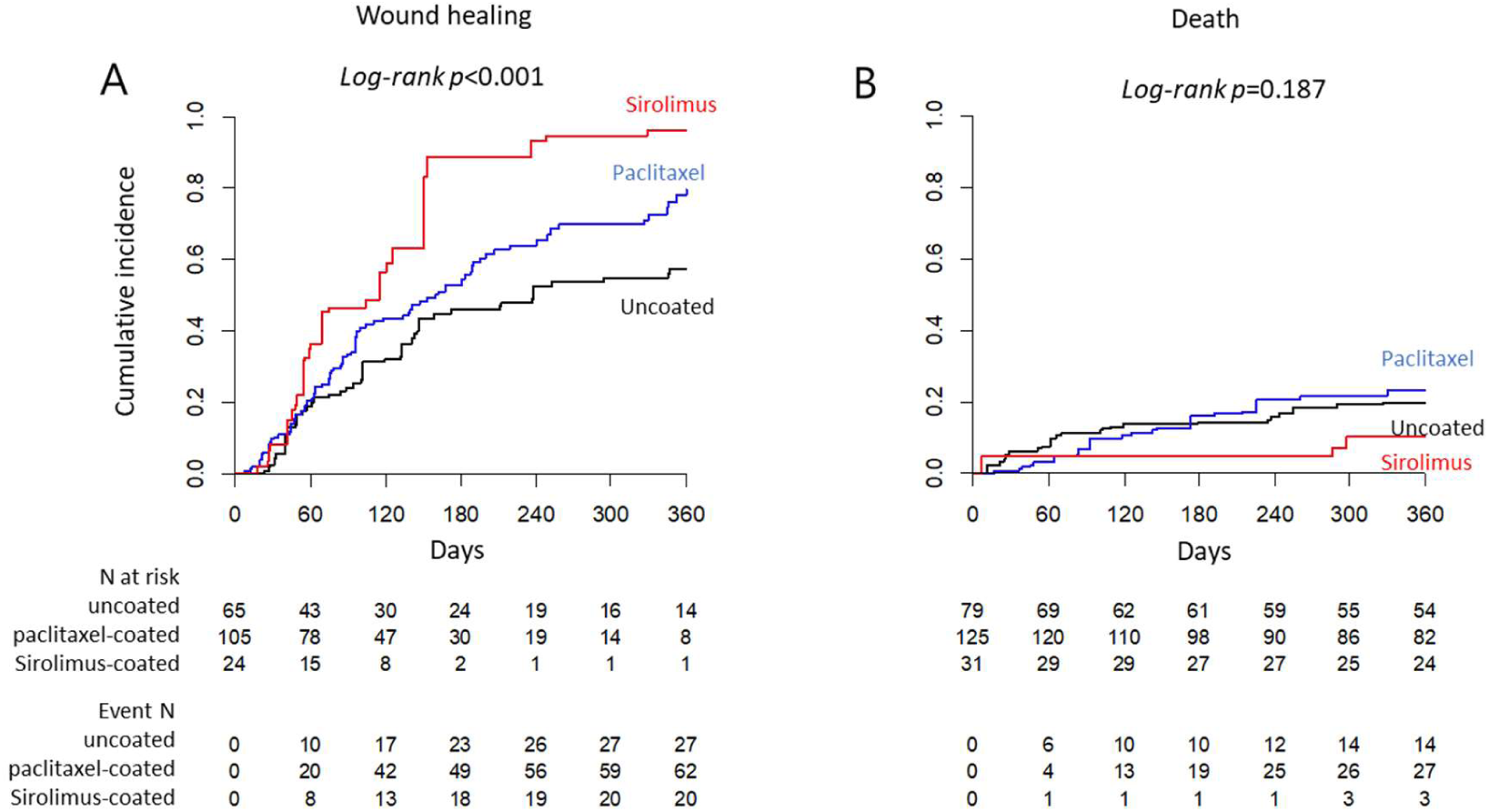
IPTW-applied cumulative incidences of wound healing and death according to drug type. In the multinomial IPTW-applied cohort of patients treated with single type of drug-coated or uncoated devices, the cumulative incidence of wound healing was highest in the sirolimus-coated device group and lowest in the uncoated device group. The cumulative incidence of death was numerically lower in the sirolimus-coated device group than in the other groups. Other clinical outcomes did not differ among groups. The red line indicates the sirolimus-coated device group, the blue line indicates the paclitaxel-coated device group and the black line indicates the uncoated device group. IPTW, inverse probability of treatment weighting.

In multinomial IPTW-applied multivariable CPH models, sirolimus-coated devices were associated with a higher probability of wound healing than both paclitaxel-coated and uncoated devices, whereas paclitaxel-coated devices were not associated with more improved wound healing than uncoated devices. Sirolimus-coated balloons were associated with lower mortality than paclitaxel-coated devices and showed a non-significant trend towards lower mortality compared with uncoated devices. In contrast, mortality was comparable between paclitaxel-coated and uncoated devices (Figure 4).

**Figure 4.**
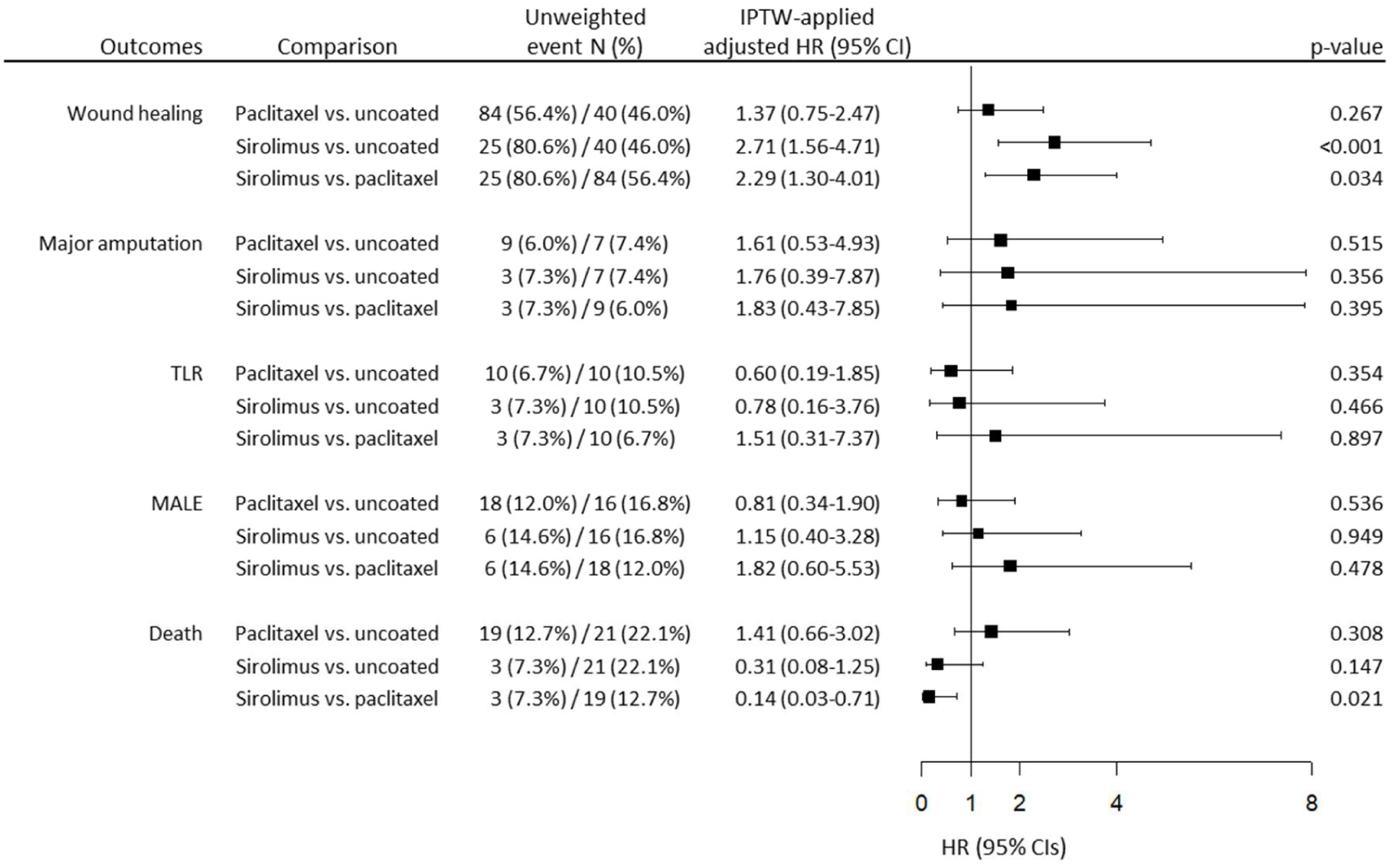
Risks of clinical events according to drug types in multinomial IPTW-applied multivariable CPH models. In the multinomial IPTW-applied cohort of patients treated with single type of drug-coated or uncoated devices, the probability of wound healing was higher in the sirolimus-coated device group than in both the uncoated and paclitaxel-coated groups, while it did not differ significantly between the paclitaxel-coated and uncoated device group. Sirolimus-coated devices were associated with a lower risk of death compared with paclitaxel-coated devices, whereas the risk of death did not differ in the other comparisons. The risks of major amputation, cdTLR and MALE did not differ among the 3 groups. IPTW, inverse probability of treatment weighting; CPH, Cox proportional hazard; cdTLR, clinically-driven target lesion revascularization; MALE, major adverse limb event.

In the subgroup without stent implantation, IPTW balanced most baseline characteristics between the DCB and PB groups, although residual differences remained in smoking, history of stroke, cholesterol levels, and lesion complexity (Table S2, Figure S5). IPTW-applied multivariable CPH models showed that DCB use was associated with more frequent wound healing and marginally lower mortality than PB angioplasty (Figure 5). In the subgroup treated without DCB, IPTW also improved balance between groups, with remaining greater lesion complexity in the DES group (Table S3, Figure S6). IPTW-applied multivariable CPH models showed that DES use was marginally associated with a higher wound healing rate than PB angioplasty with provisional BNS implantation but was not associated with other clinical outcomes (Figure 5).

**Figure 5.**
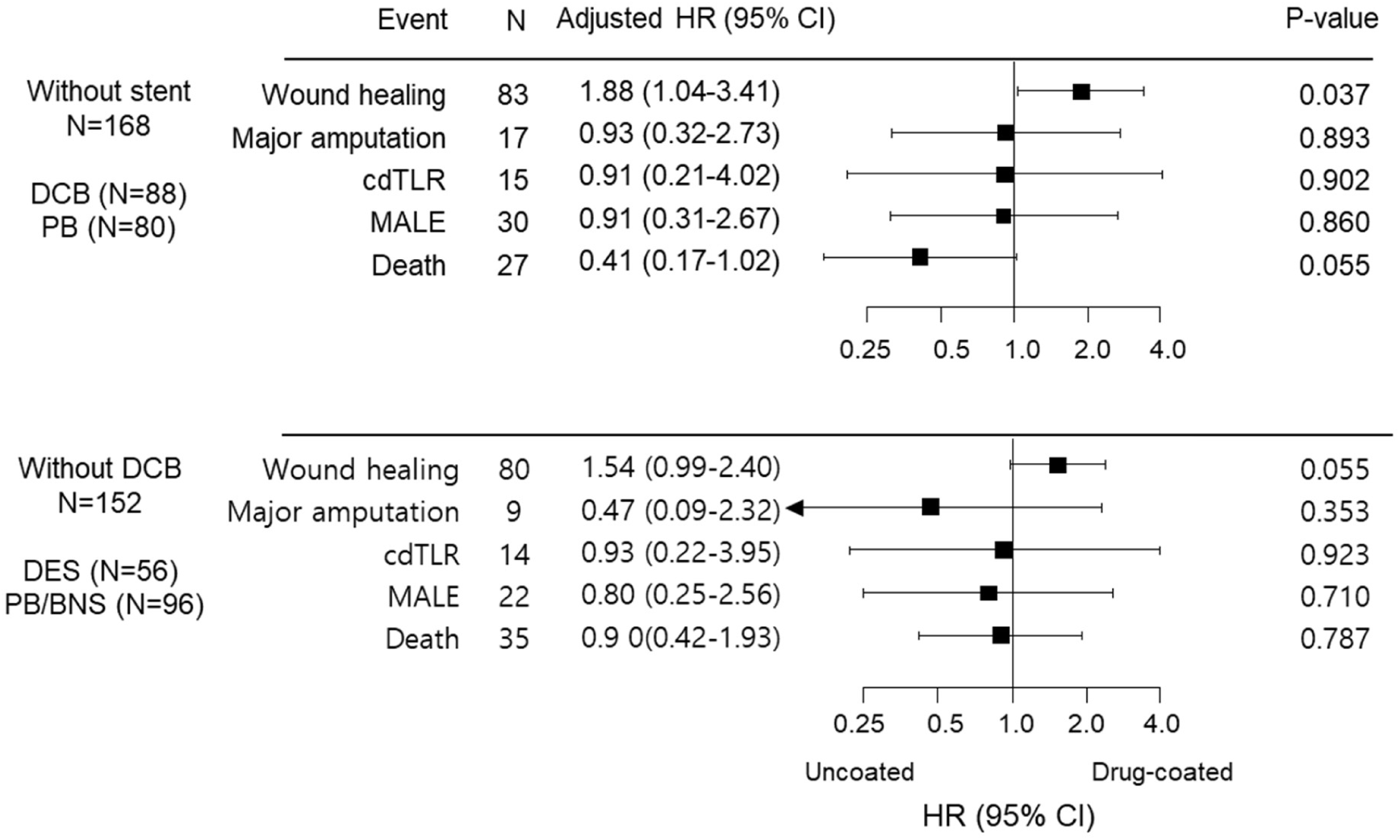
Risks of clinical events in patients treated without stents and in those treated without drug-coated balloons (DCBs). In the IPTW-applied cohorts of patients treated without stent, the wound healing rate was significantly higher in the drug-coated device group, whereas the risk of death was marginally lower in the drug-coated device group than in the uncoated device group. In the IPTW-applied cohorts of patients treated without DCBs, the wound healing rate was marginally higher in the drug-coated device group, whereas no other clinical outcomes differed significantly between the groups. * Unweighted numbers of events are shown. IPTW, inverse probability of treatment weighting; DCB, drug-coated balloon; PB, plain balloon; DES, drug-eluting stent; BNS, bare nitinol stent; HR, hazard ratio; CI, confidence interval; cdTLR, clinically-driven target vessel revascularization; MALE, major adverse limb event.

## Discussion

In this study, EVT using drug-coated devices was not associated with an increased risk of death in real-world patients with CLTI over 1 year. Drug-coated devices, including DCB and DES were associated with improved wound healing, but not with lower risks of major amputation, TLR, or MALE. In the drug-specific analysis, SCB showed the most favorable wound healing profile and was associated with lower mortality compared with paclitaxel-coated devices, whereas paclitaxel-coated and uncoated devices showed comparable mortality. These findings suggest that in contemporary CLTI practice, drug-coated devices may improve wound healing without an apparent early safety penalty.

These findings should be interpreted in the context of recent randomized evidence. SWEDEPAD 1 showed no clinical benefit of paclitaxel-coated devices in patients with CLTI over 5 years, while SWEDEPAD 2 raised safety concerns by showing higher long-term mortality in claudicants treated with paclitaxel-coated devices.^3,4^ Our study does not contradict the neutral effect of drug-coated devices on major amputation, cdTLR, or MALE. However, it adds a clinically important perspective of drug-coated devices by showing improved wound healing, which may be a more sensitive short-term endpoint than major amputation in a CLTI cohort.

Major amputation is determined not only by vascular patency but also by multiple factors including wound burden, infection, microcirculation, frailty, renal disease, and socioeconomic status.^12,13^ CdTLR also cannot capture all losses of patency because of silent restenosis and clinical decisions to defer repeat revascularization. Therefore, an EVT strategy designed to improve early patency may more readily translate into improved wound healing without producing any measurable reduction in major amputation or cdTLR in a modest-sized observational cohort as ours. This interpretation is biologically plausible. Most wound healing is achieved within 2-4 months, and restenosis or recoil after plain balloon angioplasty may compromise sustained inline flow during this period required for tissue repair.^5,14^ Because both DCB and DES are designed to enhance patency within the first year by suppressing neointimal hyperplasia, their benefits may be more relevant to wound healing than to longer-term patency or major amputation. Improved wound healing without lower cdTLR may also reflect the inability of cdTLR to capture silent restenosis and deferred or delayed procedures.^11,15^

No excess mortality signal associated with drug-coated devices, particularly paclitaxel-coated devices was observed in this study. Mortality concerns were first raised by Katsanos et al. in a meta-analysis including 28 randomized controlled trials with paclitaxel-coated devices^16^ and subsequently re-emerged in SWEDEPAD 2.^4^ However, most patients included in that meta-analysis were claudicants (89%), and SWEDEPAD 2 exclusively included claudicants. In contrast, excess mortality with paclitaxel-coated devices have not been observed in studies exclusively including patients with CLTI. Furthermore, mortality differences in the previous studies became apparent after substantially longer follow-up than the 1-year observational period of the present study. Further studies are required to clarify the long-term safety of paclitaxel-coated devices in patients with CLTI.

The favorable outcomes observed with SCB are particularly hypothesis-generating and are consistent with emerging randomized evidence for limus-based balloon technology. In the SIRONA trial, SCB showed non-inferior 12-month clinical outcomes compared with PCB in patients with femoropopliteal disease, supporting SCB as an effective alternative to PCB, although >90% of the included patients were claudicants.^8^ More recently, the SirPAD trial showed that SCB reduced 1-year MALE compared with uncoated PB in patients with infrainguinal PAD, of whom, 35% had CLTI and 30% had infrapopliteal target lesions.^6^ In our exclusively CLTI cohort, SCB was associated with better wound healing and lower mortality compared with paclitaxel-coated devices, most of which were PCBs (68.9%), suggesting clinically relevant advantages of SCB-based angioplasty over PCB-based angioplasty in CLTI.

Both sirolimus and paclitaxel inhibit smooth muscle cell proliferation by interfering cell cycle progression, although through different mechanisms.^17^ A potential advantage of SCB over PCB may be explained by their differences in drug delivery rather than by antiproliferative potency. Paclitaxel is highly hydrophobic, crystalline, and lipid-soluble. These features enable its effective tissue delivery, but also accompany geographically uneven burst-delivery pattern with a higher tendency for particulate drug emboli.^18^ In contrast, sirolimus-coated devices typically require phospholipid-encapsulated nanocarrier to deliver more hydrophilic sirolimus to the smooth muscle layer.^19^ In the SELUTION SLR platform used in this study, the carriers remain in tissue for 3 months, providing prolonged antiproliferative effects, more homogeneous drug distribution and less particulate material to embolize.^20^

No clinical studies have yet demonstrated outcome differences between PCB and SCB in head-to-head comparisons in CLTI patients. However, early evidence suggests that SCB may be less prone to the slow-flow phenomenon, which is associated with worse limb outcomes in CLTI, because it produces fewer particulate emboli than PCB.^21,22^ Although the clinical significance of drug embolization during DCB angioplasty remains uncertain and contemporary evidence has shown reassuring long-term safety regarding drug embolization,^23^ differences in the drug delivery and particulate embolization may have contributed to the wound healing difference observed in our study. Further randomized studies are required to establish whether SCB provides clinical advantages over PCB in CLTI.

This study has limitations. First, as a single-center observational study, it is susceptible to referral bias, and the observed associations cannot establish causality. Although IPTW was applied to balance the baseline differences and multivariable models were employed to further adjust for residual imbalance, remaining confounding cannot be excluded. Second, the number of patients and clinical events were modest and follow-up was limited to 1 year, which may have been insufficient to detect clinical outcome differences, in particular, mortality difference. Third, systematic post-procedural imaging surveillance was unavailable beyond 1 month; therefore, the association between patency and outcomes could not be directly assessed. Fourth, patients requiring urgent EVT or those with advanced kidney disease were transferred to the vascular hub, potentially limiting generalizability of the results to non-elective high-risk CLTI populations.

Nevertheless, our study has several strengths. It reflects an all-comer real-world CLTI population, which had infrapopliteal lesions in 61%, CTO lesions in 88%, and lesion length of 165-240 mm, representing lesion characteristics of typical real-world CLTI patients undergoing EVT.^24,25^ The study also assessed wound healing, a highly relevant short-term endpoint for patients with CLTI that is seldom investigated in randomized trials. In addition, comparisons among paclitaxel-coated, sirolimus-coated, and uncoated devices were included, which has not yet been reported in an exclusively CLTI population. Unlike SWEDEPAD 1, we also included subgroup analyses excluding stent to compare DCB with PB and excluding DCB to compare DES with PB plus provisional BNS.

In this real-world cohort of patients with CLTI undergoing infrainguinal EVT, drug-coated device use was not associated with increased adjusted 1-year mortality and was associated with improved wound healing. Exploratory drug-specific analyses suggested favorable wound-healing outcomes with sirolimus-coated balloons, with a lower observed mortality signal compared with paclitaxel-coated devices. Given the observational design, modest event numbers, and small sirolimus subgroup, these findings should be considered hypothesis-generating. Further adequately powered comparative studies should evaluate drug-coated technologies in CLTI using wound healing and other patient-relevant outcomes.

## Data Availability

The data are NHS service data and cannot be shared.

## Nonstandard Abbreviations and Acronyms

CLTI: Chronic limb-threatening ischemia;
PAD: Peripheral artery disease;
EVT: Endovascular therapy;
DCB: Drug-coated balloon;
DES: Drug-eluting stent;
eGFR: Estimated glomerular filtration rate;
eABPI: Estimated ankle-brachial pressure index;
PB: Plain balloon;
PCB: Paclitaxel-coated balloon;
SCB: Sirolimus-coated balloon;
PES: Paclitaxel-eluting stent;
BNS: Bare-nitinol stent;
cdTLR: Clinically-driven target lesion revascularization;
MALE: Major adverse limb event;
IPTW: Inverse probability of treatment weighting;
CTO: Chronic total occlusion;
SMD: Standard mean difference;
CPH: Cox proportional hazard model;
Non-HDL-C: Non-high-density lipoprotein cholesterol

## Acknowledgements

We thank to all staff in the vascular department and radiology department at the East Surrey Hospital, Surrey and Sussex Healthcare NHS Trust, for their supports and assistance with data collection.

